# The role of airborne transmission in a large single source outbreak of SARS-CoV-2 in a Belgian nursing home in 2020

**DOI:** 10.1101/2021.12.17.21267362

**Authors:** Bea Vuylsteke, Lize Cuypers, Guy Baele, Marianne Stranger, Sarah Lima Paralovo, Emmanuel André, Joke Dirks, Piet Maes, Marie Laga

## Abstract

**Objectives:** To better understand the conditions which have led to one of the largest COVID-19 outbreaks in Belgian nursing homes in 2020.

**Setting:** A nursing home in Flanders, Belgium, which experienced a massive outbreak of COVID-19 after a cultural event. An external volunteer who dressed as a legendary figure visited consecutively the 4 living units and tested positive for SARS-CoV-2 the next day. Within days, residents started to display symptoms and the outbreak spread rapidly within the nursing home.

**Methods:** We interviewed key informants and collected standardized data from all residents retrospectively. A batch of 115 positive samples with a Ct value of <37 by qRT-PCR were analysed using whole-genome sequencing. Six months after the outbreak, ventilation assessment of gathering rooms in the nursing home was done using a tracer gas test with calibrated CO_2_ sensors.

**Results:** Timeline of diagnoses and symptom onsets clearly pointed to the cultural event as the start of the outbreak, with the volunteer as index case. The genotyping of positive samples depicted the presence of one large cluster, suggesting a single source outbreak.

The global attack rate among residents was 77% with a significant association between infection and presence at the event. Known risk factors such as short distance to or physical contact with the volunteer, and wearing of a mask during the event were not associated with early infection. The ventilation assessment showed a high background average CO_2_ level in four main rooms varying from 657 ppm to 846 ppm.

**Conclusions:** Our investigation shows a rapid and widespread single source outbreak of SARS-CoV-2 in a nursing home, in which airborne transmission was the most plausible explanation for the massive intra-facility spread. Our results underscore the importance of ventilation and air quality for the prevention of future outbreaks in closed facilities.

## Introduction

Since the beginning of the ongoing COVID-19 pandemic, numerous outbreaks of SARS-CoV-2 have been reported in nursing homes for elderly people, with devastating consequences (1).

In Belgium, a first COVID-19 wave occurred from March until June 2020, and was followed by a second wave from September 2020 till February 2021. At an early stage, infection prevention and control measures were scaled up in nursing homes, including banning of family visits and regular testing of residents and staff. By June 21, 2020, 90% of Belgian nursing homes reported at least one case of SARS-CoV-2 infection among residents, and 40% reported at least ten cases (2). More than half of all COVID-19 related deaths in 2020 in Belgium were linked to nursing homes (3). Residents and staff of nursing homes were the first priority group in the Belgian vaccination campaign. By March 2021, vaccination coverage among residents of nursing homes had reached 89.4%, and serious COVID-19 outbreaks in nursing homes became rare (4).

The nursing home “Z” in Flanders, Belgium, had been spared from a COVID-19 outbreak until late 2020. In October, 11 residents tested positive for SARS-CoV-2 (20B lineage) during a routine screening. All remained asymptomatic, and by mid-November the quarantine of the positive residents was lifted. In December 2020, after 9 months of strict lockdown, a first social event was organized in “Z”. An external volunteer who dressed as a legendary figure visited the nursing home to bring gifts to the residents. The day after, the volunteer started to develop symptoms and tested positive for SARS-CoV-2. Within days, residents started to display symptoms and the outbreak spread rapidly within the nursing home.

To better understand the conditions which have led to what turned out to be one of the largest outbreaks in Belgian nursing homes in 2020, a detailed retrospective outbreak investigation was conducted. In this paper we describe the spread of SARS-CoV-2 among the residents and explore potential risk factors for infection.

## Methods

### Setting and data collection

The nursing home “Z” is part of an international independent provider of senior care and has a maximum capacity of 179 residents, who live in single or double rooms. The facility is structured in four separate living units (hereafter called “A”, “B”, “C” and “D”), with each of them containing separate catering and gathering spaces, as well as a large common cafeteria, which was accessible to all residents and visitors before the COVID-19 pandemic.

The investigators (BV & ML) visited “Z” during the last days of the outbreak, and interviewed two key informants. Based on this interview, a line listing using a spreadsheet software program was designed to contain basic information on all residents. This information included selected demographic characteristics, SARS-CoV-2 RT-PCR tests and results, information on symptoms and outcome if tested positive, and some behaviour characteristics during the event. The coordinating physician (JD) and an occupational therapist who was present during the event collected the data retrospectively. Data were pseudonymised before transferring to the Institute of Tropical Medicine.

### Genome analysis of the positive samples

A batch of 115 positive samples with a Ct value of <37 by qRT-PCR were analysed using whole-genome sequencing. RNA extraction was performed using the QIAampViral RNA extraction kit, while the genomes were amplified using the ARTIC network V3 protocol. After clean-up of the amplicons, libraries were prepared using the ligation sequencing kit (LSK109) from Oxford Nanopore Technologies. Subsequently, the libraries were quantified, and sequencing was performed on a GridION using R9.4.1 flow cells. Sequencing runs were processed using the ARTIC analysis pipeline and custom scripts. Full-length genome sequences accompanied with metadata were submitted to GISAID (accession numbers EPI_ISL_738361 to 412, EPI_ISL_738414 to 418, EPI_ISL_738420 to 424, EPI_ISL_738426 to 435, EPI_ISL_738437 to 441, EPI_ISL_738443 to 446, EPI_ISL_738449 to 452, EPI_ISL_738454, EPI_ISL_738456 to 460, EPI_ISL_738462 to 466, EPI_ISL_738468 to 469, EPI_ISL_738471, EPI_ISL_738473 to 474, EPI_ISL_738478 to 479, EPI_ISL_738481, EPI_ISL_738484 to 485, EPI_ISL_738487, EPI_ISL_738490 to 491, EPI_ISL_738493, EPI_ISL_738495 to 497, EPI_ISL_738499, and EPI_ISL_745222). SARS-CoV-2 lineage assignments were derived using the Pangolin tool as well as classification by Nextclade (https://clades.nextstrain.org) (5,6).

The sequences were aligned with MAFFT v7.310, and phylogenetic analysis was conducted with IQ-TREE v1.6.12, accompanying the set of 115 newly obtained genomes with a representative set of 1467 international SARS-CoV-2 sequences available on GISAID at the time of the analysis selected by using NextStrain (7).

### Quality of ventilation

At the request of the Flemish Government, a team from VITO conducted an assessment of the ventilation conditions of the nursing home in July 2021, when the COVID-19 incidence lowered significantly and nursing homes were accessible again for visitors not related to occupants. In total, four large rooms where the residents frequently gather (for eating, watching TV and socializing in general) were assessed via tracer gas test for ventilation rates measurements (8). In each of the assessed rooms, calibrated CO_2_ sensors with automatic logging and a resolution of 1 minute were distributed across the space before the tracer gas injection, in order to determine the CO_2_ background concentration. After a stabilization period of approximately 15 minutes, the CO_2_ gas injection was initiated, at a high and controlled emission rate. The injection was stopped when a CO_2_ concentration sufficiently high was achieved, i.e. at least 1000 ppm above the (room) background. After the team exited the room, the CO_2_ sensors monitored the CO_2_ concentration decay for a total period of 40 minutes to 1 hour. The ventilation rate of each room was then derived from the rate of concentration decay monitored by the sensors. The ventilation rate was determined during normal operational conditions of each room, which implies that open (inner and outdoor) doors were not closed during the assessment.

### Statistical methods

The analysis and graphs were done using R version 4.0.2 (R Foundation for Statistical Computing, Vienna, Austria, www.R-project.org).

Odds Ratios resulting from logistic regression were calculated to measure the strength of associations in bivariate and multivariate models.

### Ethical approval

Ethical approval for the study was obtained from the Institutional Review Board of the ITM, and Ethics Committee of the Group owning the nursing home.

## Results

### Timeline of SARS-CoV-2 outbreak

The sequence of events related to the outbreak is summarized in Figure 1. All staff had tested negative on the last monthly screening four days before the outbreak. The social event, with the visit of the volunteer, toke place on day 0 (Event). The residents of Unit “D” were in quarantine on day 0, because one resident of the unit, while in hospital, had tested positive a few days before. All residents of unit “D” tested negative on day 0. One day later, the volunteer developed symptoms and tested positive for SARS-CoV-2 with Ct values < 13.5 for all three target genes categorized as “very high viral load”. A first resident (from unit “A”) started having symptoms suggestive of COVID-19 four days after the event. Selected testing was initiated, mostly guided by symptoms, followed by rounds of general testing (details see Figure 1). Because of the fear of an emerging outbreak, all residents were confined to their rooms. The infected residents were cared for in dedicated COVID-19 sections of the nursing home, and additional protective equipment for the nursing staff was made available. The proportion of positive tests was 35.6%, 42.0%, 26.4%, 16.2% and 8.8%, on respectively day 7, 10, 14, 19 and 26 after the event. On day 9 a first resident died of COVID-19. By the end of December, a total of 127 residents and 40 staff were diagnosed with SARS-CoV-2 since the beginning of the outbreak. Among the infected residents, 77 experienced COVID-19 symptoms (60.6%) and 35 died, resulting in a case fatality rate of 27.6%.

**Figure 1.**
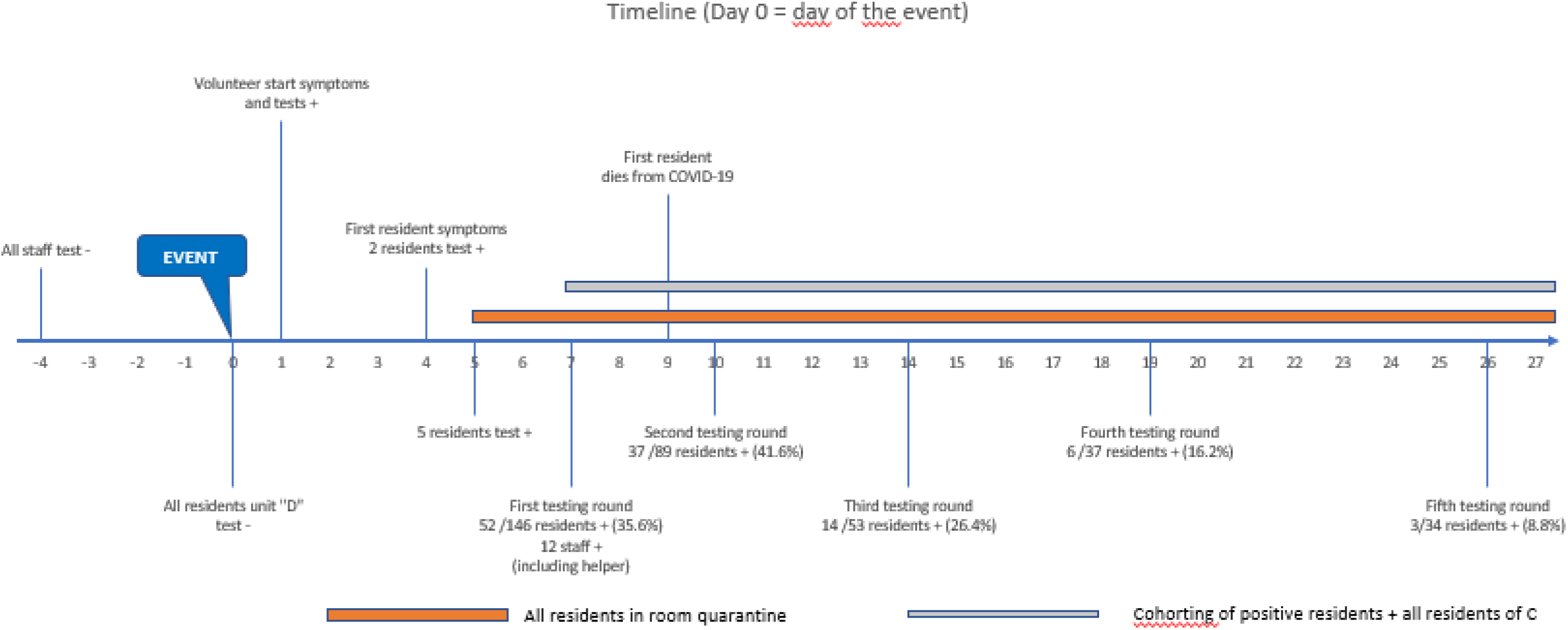
Timeline of events. Series of testing events in nursing home “Z” following the visit of an external volunteer (day 0, indicated by the Event flag).

### Detailed description of the event

An external volunteer who dressed as a legendary figure visited consecutively four rooms where residents of living units A, B and C were gathered. Three staff members, of whom two were dressed up as helpers to hand out gifts, were part of the company. Visits lasted from 15 to 30 minutes per room. Finally, the company strolled through the corridor of unit “D” whose residents remained in their rooms.

Because of confidentiality concerns, a detailed description of the event and flow of the visit is not published here, but is available upon request.

### Genotyping of the samples

The 115 samples for genotyping were collected at an early stage in the epidemic and originated from 97 residents, 17 staff and 1 (external) volunteer. Ct values were < 40 for all three target genes. All 115 SARS-CoV-2 sequences could be assigned to clade 20B according to Nextclade, while phylogenetically the lineage B.1.1 was derived. Phylogenetic inference (see Figure 2) of a large set of publicly available B.1.1 genomes clearly depicts the presence of one large cluster, embedding all sequences of the outbreak, providing evidence of one circulating strain. Except for a limited number of point mutations, sequences were found to be identical, suggesting a single source outbreak. Apart from the amino acid substitution D614G, a mutation that is prevalent in nearly all Belgian SARS-CoV-2 genomes since the start of the epidemic, no changes were reported in the Spike protein compared to Wuhan reference NC_045512 (9).

**Figure 2.**
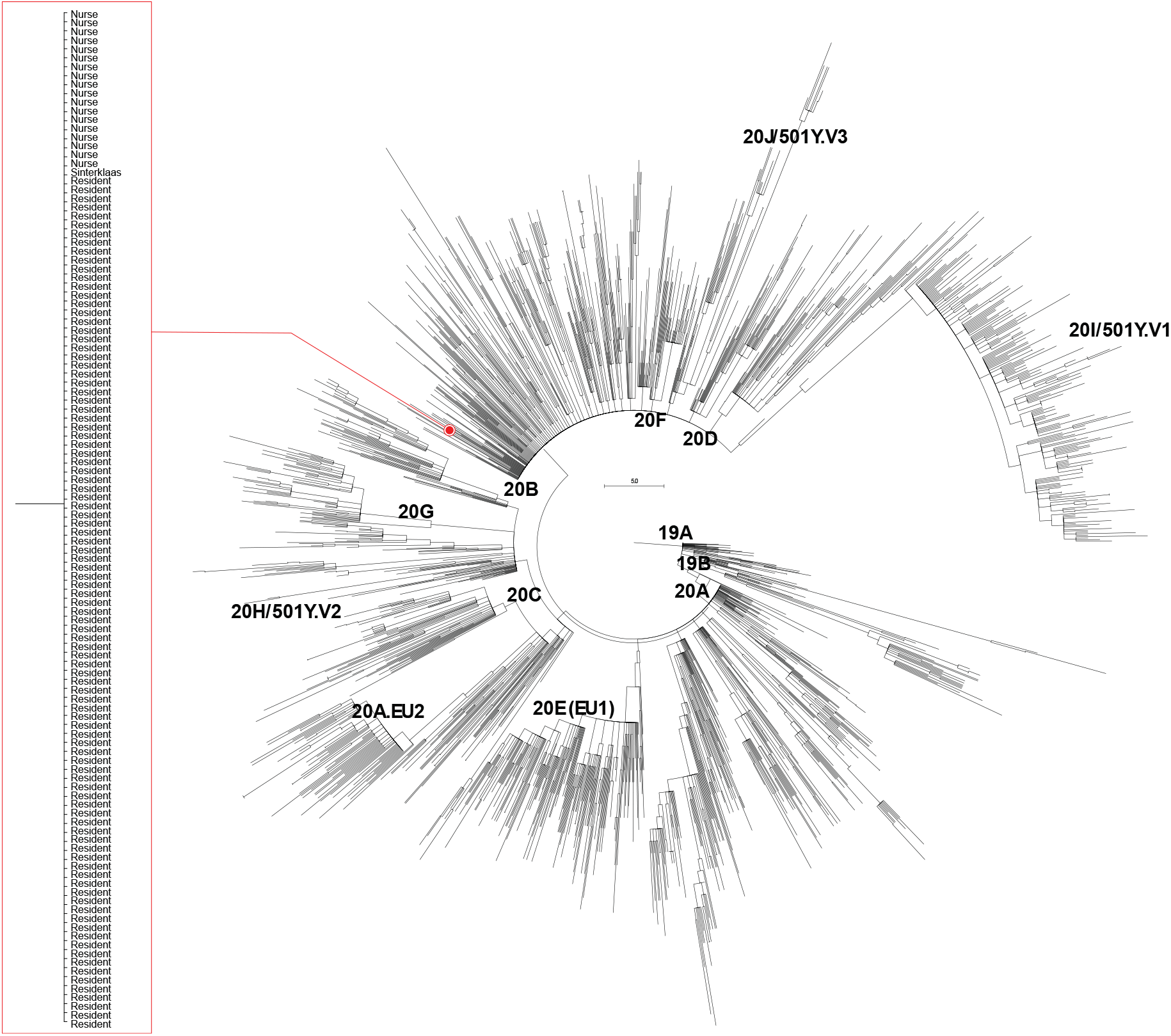
Phylogenetic tree of B.1.1. Maximum likelihood tree showing the genomes of the nursing home outbreak cluster as one large monophyletic cluster (see inset) within the SARS-CoV-2 lineage 20B cluster, confirming a single introduction event into the nursing home.

### SARS-CoV-2 Attack rates per living unit and associated characteristics among residents

By the end of the outbreak, 127 (77%) residents had tested positive, and the attack rates differed by living unit (see Figure 3). The steepest increase was seen around day 6 in the 3 units participating in the event (“A”, “B” and “C”). In unit “D” the main increase of infections was seen on day 10. The cumulative attack rate of unit “D” also remained lower: 52.5 % as compared to the other units: 84.5%, 92.1% and 77.8% in respectively living unit “A”, “B” and “C”.

**Figure 3.**
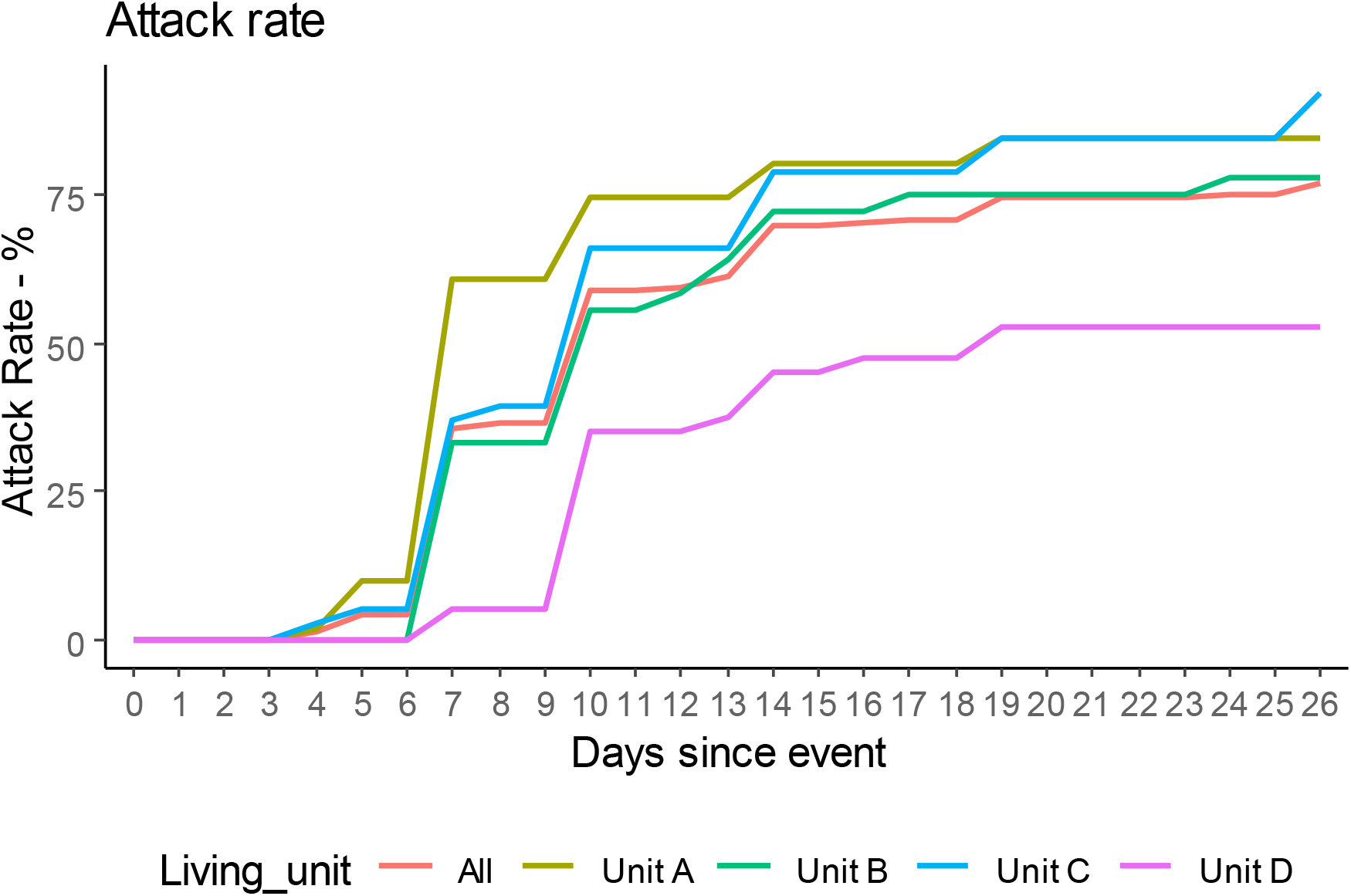
Cumulative attack rate (AR) per living unit. Units A, B and C showed similar patterns over time, whereas the severity of the outbreak in unit D was markedly lower.

At the time of the event, there were 124 women (75.2%) and 41 men (24.8%) residing in “Z”. The median age was 87 years old, IQR 82-91, with a minimum of 62 and a maximum of 100 years. A total of 97 residents out of the 165 (88.7%) participated in the event. Attack rates were higher among female residents, and among residents older than 80 years, albeit not significantly. Nine residents who experienced an asymptomatic SARS-CoV-2 infection in October 2020 did not have a lower attack rate. Participation in the event was the only variable significantly associated with a higher attack rate (Table 1). Living unit was not included in the multivariable analysis, because of collinearity with presence at the event (none of the residents of unit “D” were present in a gathering room).

**Table 1:** Attack rate (AR) and association with selected characteristics among residents.

|  |  |  | Logistic regression |  | Multivariable logistic regression |  |
| --- | --- | --- | --- | --- | --- | --- |
|  | N | AR (%) | OR | 95% CI | AOR | 95% CI |
| Sex |  |  |  |  |  |  |
| M | 41 | 73.17 | 1 |  |  |  |
| F | 124 | 78.23 | 1.32 | (0.57-2.92) | 1.32 | (0.5-3.36) |
| Age |  |  |  |  |  |  |
| <80 | 28 | 64.29 | 1 |  |  |  |
| 80-89 | 85 | 78.82 | 2.07 | (0.8-5.23) | 1.66 | (0.56-4.75) |
| 90+ | 52 | 80.77 | 2.33 | (0.82-6.67) | 1.97 | (0.6-6.46) |
| Living unit |  |  |  |  |  |  |
| "A" | 51 | 84.31 | 1 |  |  |  |
| "B" | 36 | 77.78 | 0.65 | (0.21-1.96) | (-) | (-) |
| "C" | 38 | 92.11 | 2.17 | (0.58-10.47) | (-) | (-) |
| "D" | 40 | 52.50 | 0.21 | (0.07-0.53) | (-) | (-) |
| Tested PCR positive for SARS-CoV-2 in October 2020 |  |  |  |  |  |  |
| No | 149 | 81.88 | 1 |  |  |  |
| Yes | 9 | 55.56 | 0.35 | (0.09-1.47) | 0.24 | (0.05-1.19) |
| Participated in the event |  |  |  |  |  |  |
| No | 68 | 60.29 | 1 |  |  |  |
| Yes | 97 | 88.66 | 5.15 | (2.38-11.79) | 5.64 | (2.34-14.63) |
| Had contact with helper |  |  |  |  |  |  |
| No | 59 | 69.49 | 1 |  |  |  |
| Yes | 106 | 81.13 | 1.89 | (0.9-3.96) | 0.94 | (0.38-2.25) |

AR: Attack Rate
OR= Odds Ratio; AOR= Adjusted Odds Ratio in multivariable logistic regression

### Timing of SARS-CoV-2 diagnoses and attendance at the event

We assumed infections detected within 8 days of the event were likely directly linked to the event and defined them as early infections. Participation in the event was associated with higher early infection rates, but some residents who had not participated in the event were also early infected. Among the 97 residents who were present at the event, 86 (88.7%) became infected with SARS-CoV-2. Of these 86 infections, 45 (52.3%) were detected within 8 days. Among the 68 residents who were not present at the event, 41 (60.3%) became infected with SARS-CoV-2. Of these 41 infections, 14 (34.1%) were detected within 8 days. More detailed information on the timing of SARS-CoV-2 diagnoses and attendance at the event by living unit is shown in Supplementary Figure 1.

### *Factors associated with early infections among attendees of the* event

Risk factors for early infection among the 97 residents who participated in the event were further explored in a nested case control analysis. Cases were residents who participated in the event and tested positive within 8 days after the event (N=45), and controls were residents who participated in the event but remained negative or tested positive after 14 days or later (N=20). Gathering room, distance to the volunteer, wearing of a mask or contact with the volunteer were not associated with early infection. Only having had contact with a helper was associated with early infection (OR = 5.86, 95%CI: 1.36 - 30.81) (Table 2)

**Table 2:** Nested case control study among residents who participated in the event.

|  | Cases | % | Controls | % | OR | 95% CI |
| --- | --- | --- | --- | --- | --- | --- |
|  | N= 45 |  | N=20 |  |  |  |
| Gathering Room |  |  |  |  |  |  |
| Multifunctional room "A" | 26 | 57.8 | 8 | 40.0 | 1 |  |
| Cafeteria | 10 | 22.2 | 7 | 35.0 | 0.44 | (0.12-1.55) |
| Eating room "C" | 2 | 4.4 | 1 | 5.0 | 0.61 | (0.05-14.26) |
| TV room "C" | 7 | 15.6 | 4 | 20.0 | 0.54 | (0.12-2.48) |
| Distance to index case |  |  |  |  |  |  |
| <2m | 9 | 20.5 | 3 | 15.0 | 1 |  |
| 2-3m | 15 | 34.1 | 4 | 20.0 | 1.25 | (0.21-7) |
| 4-5m | 15 | 34.1 | 7 | 35.0 | 0.71 | (0.13-3.33) |
| 5m+ | 5 | 11.4 | 6 | 30.0 | 0.28 | (0.04-1.54) |
| Wearing mask |  |  |  |  |  |  |
| No | 15 | 33.3 | 10 | 50.0 | 1 |  |
| Yes | 30 | 66.7 | 10 | 50.0 | 2.14 | (0.76-6.43) |
| Contact with volunteer |  |  |  |  |  |  |
| No | 34 | 75.6 | 17 | 85.0 | 1 |  |
| Yes | 10 | 22.2 | 3 | 15.0 | 1.67 | (0.44-8.15) |
| Contact with helper |  |  |  |  |  |  |
| No | 4 | 8.9 | 6 | 30.0 | 1 |  |
| Yes | 41 | 91.1 | 14 | 70.0 | 5.86 | (1.36-30.81) |

OR = Odds Ratio

### Quality of ventilation in the nursing home

The assessment of the quality of ventilation was performed in four main living rooms, including two rooms where the residents of “C” were gathered during the event. Those rooms were equipped with openable windows and exterior doors, and a mechanical ventilation system with grids above the windows and controlled air extraction. The other characterized rooms were also rooms where residents frequently gather for eating, watching TV and socializing. These rooms were used by the residents prior and right after the event. They were not equipped with ventilation facilities, only with a fan coil unit for air heating and cooling. The background average CO_2_ level in the four rooms varied from 657 ppm to 846 ppm and the maximum background concentration varied from 708 to 897 ppm. The lowest CO_2_ background levels were determined in both rooms where the residents of “C” were gathered during the outbreak event. The air change rate determined by the different CO_2_ sensors in the four rooms during the tests varied between 3.6/h and 2.9/h in both rooms with opened door and windows, and between 1.8/h and 2.3/h in the rooms without ventilation facilities.

The cafeteria where residents of “B” were gathered during the cultural event was not included in the ventilation assessment. It is the largest room with 838 m^2^, not equipped with ventilation facilities, only with a fan coil unit for air heating and cooling, which was operating at the time of the visit. The other room where residents of “A” gathered for the event were equipped with simple flow controlled mechanical ventilation, with natural air supply through ventilation grids at the windows and mechanical air exhaust. Information about the use of ventilation grids and air extraction during the event is unknown.

## Discussion

Our investigation shows a rapid and widespread outbreak of SARS-CoV-2 in a nursing home with an identical virus pointing to a single source. The virus was most likely introduced into the nursing home by the volunteer and amplified by the social event. Aerosol transmission in crowded poorly ventilated spaces is the most plausible explanation for the massive intra-facility spread.

Severe outbreaks of COVID-19 in nursing homes have been reported all over the world throughout 2020. The attack rate of 77% among 165 residents in this study is much higher than attack rates previously reported in nursing homes during the same, pre-vaccination, period. A systematic review and meta-analysis of 49 studies from four continents showed a pooled attack rate of 28% (95% CI 18-40%). When limiting the analysis to the studies reporting outbreaks from a single facility, the pooled attack rate was 45% (95% CI 32-58%) (10). This very high attack rate is worrisome. Especially when considering the many measures which were put in place to prevent the spread of Covid-19, including continuous infection control measures for staff, the ban of visitors and the implementation of room quarantine and cohorting of infected residents.

Based on the timeline of diagnoses and symptom onsets, and the significant association we found between infection and presence at the event, we can point to the cultural event as the most plausible start of the outbreak. In addition, the phylogenetic analysis showed one nearly identical viral strain pointing towards the occurrence of a single source outbreak. The volunteer was pre-symptomatic with a very high viral load on the day of the event and had contact with the four units of the nursing home during the same day. He was also the first to develop COVID-related symptoms, and therefore most likely the index case. Introduction of the virus by a staff member during the same period cannot be excluded but is unlikely, as most of the staff were confined to one unit and all of them tested negative during a routine testing four days before the event. One of the helpers of the volunteer tested positive on day 8 but started having symptoms only after a few days. It is very unlikely that this person was already infectious on the day of the event. The association we found between early infection and contact with a helper remains unexplained. The large majority of residents participating in the event had a brief contact with a helper when he/she handed out a present. In the absence of other valid hypotheses, we think this association was a random finding, also taking into account the very small number of residents who did not have a contact with a helper.

It is well established that SARS-CoV-2 is readily transmitted in indoor environments (11–13). Airborne transmission has being recognized as an important route of transmission (14,15). We have several arguments to support the dominance of airborne transmission in this outbreak. First of all, the social event was typically a crowded prolonged indoor happening in gathering rooms with no adequate ventilation. In addition, mask wearing was sub-optimal. The long beard of the volunteer made it difficult to effectively fit the surgical mask, and many residents were not wearing masks. Furthermore, we could not demonstrate the role of direct contact or droplets in this outbreak. Indeed, among those present at the event, the case control study showed no association between early infection and distance to or contact with the volunteer. Finally, the fact that some residents not present at the event were early infected may seem puzzling, but can also point to long-range aerosol transmission. The gathering rooms were connected by corridors where residents stroll. Moreover, some residents who did not participate in the event took a coffee in the same room shortly after the event. The virus may have spread through aerosols as long as three hours after the event (16).

Superspreading events, in which many people are infected at once, typically by a single individual, are a now-familiar feature of the COVID-19 pandemic (12). Detailed analysis of previous superspreading events confirm aerosols as the main driver of large outbreaks on cruise ships, in choirs or slaughterhouses (12). Biological factors may also play an important role in superspreading, such as the variability in viral shedding (17). Some subjects are able to infect a large number of people while others have a lower ability to spread the virus. The PCR test of the volunteer as presumed index case had a very low Ct value (<14 for all three targets) suggesting high viral spreading and the volunteer as potential hyperspreader.

Taken the previous into account, ventilation may play a key role in the control of SARS-CoV-2. Our measurements indicate a rather poor ventilation quality in the nursing home. Background CO_2_ concentrations in the assessed rooms (average 657 ppm to 846 ppm) were considerably higher than would be expected for a well-ventilated room with low or no occupancy. Taking into account the rural environment of the nursing home, one would expect an average outdoor CO_2_ concentration between 400-450 ppm, and thus expecting an equal background level inside the rooms. In addition, measurements took place in summer time, with exterior doors and windows open, probably resulting in lower CO_2_ concentrations than the concentration at the time of the event. The mean outside temperature on the day of the event was 5.5°C (min 1.6 – max 7.5 °C), therefore windows and ventilation grids were probably closed during the event.

(https://www.meteobelgie.be/klimatologie/waarnemingen-en-analyses/jaar-2020/2290-waarnemingen-december-2020).

The Belgian national guidelines for COVID-19 prevention recommend indoor CO_2_-levels of maximum 900 ppm (or a fresh air supply of 40 m^3^/h per person) (https://www.info-coronavirus.be/en/ventilation/). In addition, none of the rooms assessed reached the latest recommendations from the Federation of European Heating, Ventilation and Air Conditioning Associations (REHVA) nor the ventilation rates suggested in the interim guide on COVID-19 infection prevention and control during health care of the World Health Organization (WHO) (18,19).

On the other hand, if heating, ventilation and air conditioning systems are not correctly used, they may contribute to the transmission/spreading of airborne diseases as demonstrated in the past for SARS (20). In the largest room during the event, the only air treatment system was a fan coil unit, a device that uses a coil and a fan to heat or cool a room without outdoor air supply. Indoor air moves over the coil, which heats or cools the air before pushing it back out into the room. To prevent the spread of an airborne virus, the use of such installations is highly dissuaded since instead of diluting indoor concentration, they distribute potential infectious particles in a room.

This investigation was limited by its retrospective design. Data on the event, and the behaviour of the residents were collected by, and through interviews with staff members. Because of the dramatic nature of the outbreak, with high morbidity and mortality and negative reports of the mainstream media, recall bias and underreporting of risky behaviours such as close contacts with the index case, cannot be excluded.

In conclusion, we described a single source outbreak of SARS-CoV-2 in a nursing home in Belgium, in which airborne transmission was the most plausible explanation for the massive intra-facility spread. Our results underscore the importance of ventilation and air quality for the prevention of future outbreaks of SARS-CoV-2 variants in closed facilities, even in the era of high vaccination coverage.

## Data Availability

No additional data is available

## Supplementary materials

**Supplementary Figure 1:**
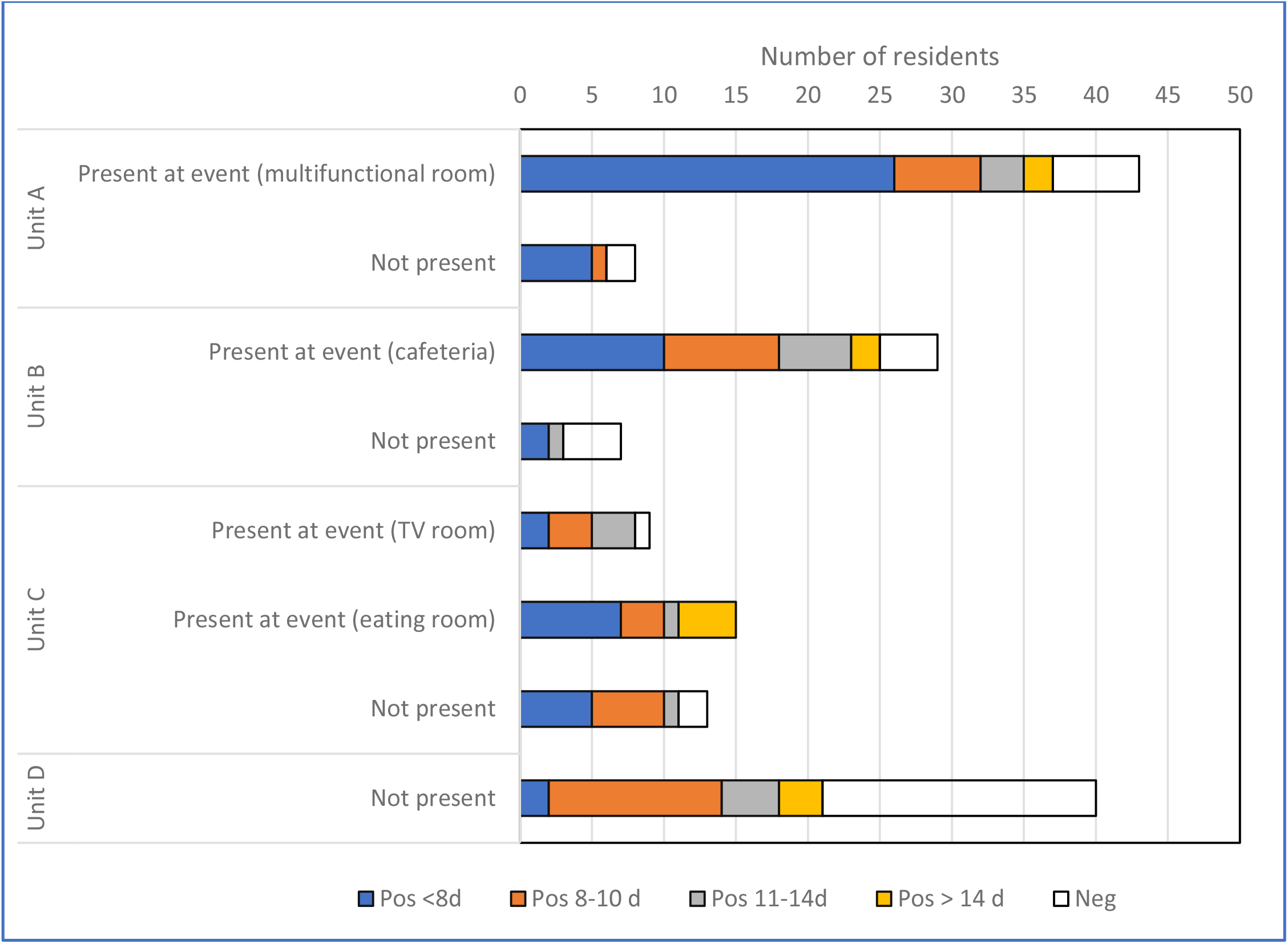
Timing of SARS-CoV-2 diagnoses by living unit and presence at event. Timing of positive tests: early infections (Pos < 8 d); tested positive between 8 and 10 days after the event (Pos 8-10d); tested positive between 11 and 14 days after the event (Pos 11-14d); tested positive more than 14 days after the event (Pos > 14d) and remained negative (Neg).

## References

1. Thompson DC, Barbu MG, Beiu C, Popa LG, Mihai MM, Berteanu M, et al. The Impact of COVID-19 Pandemic on Long-Term Care Facilities Worldwide: An Overview on International Issues [Internet]. Vol. 2020, BioMed Research International. Hindawi Limited; 2020 [cited 2021 May 20]. Available from: https://pubmed.ncbi.nlm.nih.gov/33204723/

2. Callies M, Int Panis L, Dequeker S, Latour K, Rebolledo Gonzalez J, Vandael E. COVID-19 surveillance in woonzorgcentra [Internet]. Brussels; 2021. Available from: https://covid-19.sciensano.be/sites/default/files/Covid19/COVID-19_Surveillance_WZC.pdf

3. Sciensano. COVID-19 Epidemiologisch bulletin van 2 oktober 2021 [Internet]. Brussels; 2021. Available from: https://covid-19.sciensano.be/sites/default/files/Covid19/Meestrecenteupdate.pdf

4. Catteau L, Haarhuis F, Dequeker S, Vandaele E, Stouten V, Lizroth A, et al. Surveillance van de COVID-19 vaccinatie in Belgische woonzorgcentra -Resultaten van de gegevensverzameling tot en met 24 maart 2021. [Internet]. Brussels; 2021. Available from: https://covid-19.sciensano.be/sites/default/files/Covid19/COVID-19_THEMATICREPORT_SURVEILLANCEVANDEVACCINATIEINBELGISCHEWOONZORGCENTRA.pdf

5. Rambaut A, Holmes E, O’Toole Á, Hill V, McCrone J, Ruis C, et al. A dynamic nomenclature proposal for SARS-CoV-2 lineages to assist genomic epidemiology. Nat Microbiol [Internet]. 2020 Nov 1 [cited 2021 Oct 27];5(11):1403–7. Available from: https://pubmed.ncbi.nlm.nih.gov/32669681/

6. O’Toole Á, Scher E, Underwood A, Jackson B, Hill V, McCrone JT, et al. Assignment of epidemiological lineages in an emerging pandemic using the pangolin tool. Virus Evol [Internet]. 2021 Oct 6 [cited 2021 Oct 13];7(2). Available from: https://academic.oup.com/ve/article/7/2/veab064/6315289

7. Katoh K, Standley D. MAFFT multiple sequence alignment software version 7: improvements in performance and usability. Mol Biol Evol [Internet]. 2013 Apr [cited 2021 Oct 27];30(4):772–80. Available from: https://pubmed.ncbi.nlm.nih.gov/23329690/

8. Sherman MH. Tracer-gas techniques for measuring ventilation in a single zone. Build Environ. 1990 Jan 1;25(4):365–74.

9. Wawina-Bokalanga T, Martí-Carreras J, Vanmechelen B, Bloemen M, Wollants E, Laenen L, et al. Genetic diversity and evolution of SARS-CoV-2 in Belgium during the first wave outbreak. bioRxiv [Internet]. 2021 Jun 29 [cited 2021 Aug 25];2021.06.29.450330. Available from: https://www.biorxiv.org/content/10.1101/2021.06.29.450330v1

10. Hashan MR, Smoll N, King C, Ockenden-Muldoon H, Walker J, Wattiaux A, et al. Epidemiology and clinical features of COVID-19 outbreaks in aged care facilities: A systematic review and meta-analysis. EClinicalMedicine [Internet]. 2021;33:100771. Available from: https://doi.org/10.1016/j.eclinm.2021.100771

11. Gabutti G, D’anchera E, De Motoli F, Savio M, Stefanati A. The epidemiological characteristics of the covid-19 pandemic in europe: Focus on Italy [Internet]. Vol. 18, International Journal of Environmental Research and Public Health. MDPI AG; 2021 [cited 2021 May 20]. p. 1–14. Available from: /pmc/articles/PMC8000566/

12. Lewis D. Superspreading drives the COVID pandemic - and could help to tame it. Nature [Internet]. 2021 Feb 1 [cited 2021 May 21];590(7847):544–6. Available from: https://pubmed.ncbi.nlm.nih.gov/33623168/

13. Goodwin L, Hayward T, Krishan P, Nolan G, Nundy M, Ostrishko K, et al. Which factors influence the extent of indoor transmission of SARS-CoV-2? A rapid evidence review. J Glob Health [Internet]. 2021 Apr 3 [cited 2021 May 20];11. Available from: /pmc/articles/PMC8021073/

14. Morawska L, Milton DK. It Is Time to Address Airborne Transmission of Coronavirus Disease 2019 (COVID-19) [Internet]. Vol. 71, Clinical Infectious Diseases. Oxford University Press; 2020 [cited 2021 May 21]. p. 2311–3. Available from: https://pubmed.ncbi.nlm.nih.gov/32628269/

15. Greenhalgh T, Jimenez JL, Prather KA, Tufekci Z, Fisman D, Schooley R. Ten scientific reasons in support of airborne transmission of SARS-CoV-2. Lancet (London, England) [Internet]. 2021 Apr 15 [cited 2021 May 21];397(10285). Available from: http://www.ncbi.nlm.nih.gov/pubmed/33865497

16. van Doremalen N, Bushmaker T, Morris D, Holbrook M, Gamble A, Williamson B, et al. Aerosol and Surface Stability of SARS-CoV-2 as Compared with SARS-CoV-1. N Engl J Med [Internet]. 2020 Apr 16 [cited 2021 Sep 30];382(16):1564–7. Available from: https://pubmed.ncbi.nlm.nih.gov/32182409/

17. Al-Tawfiq JA, Rodriguez-Morales AJ. Super-spreading events and contribution to transmission of MERS, SARS, and SARS-CoV-2 (COVID-19). J Hosp Infect [Internet]. 2020 Jun 1 [cited 2021 May 20];105(2):111–2. Available from: /pmc/articles/PMC7194732/

18. REVHA. How to operate HVAC and other building service systems to prevent the spread of the coronavirus (SARS-CoV-2) disease (COVID-19) in workplaces. Federation of European Heating, Ventilation and Air Conditioning Associations; 2020.

19. World Health Organization. Infection prevention and control during health care when coronavirus disease (COVID-19) is suspected or confirmed [Internet]. 2021. Available from: https://www.who.int/publications/i/item/WHO-2019-nCoV-IPC-2021.1

20. Correia G, Rodrigues L, Gameiro da Silva M, Gonçalves T. Airborne route and bad use of ventilation systems as non-negligible factors in SARS-CoV-2 transmission. Med Hypotheses [Internet]. 2020 Aug 1 [cited 2021 Jun 16];141. Available from: https://pubmed.ncbi.nlm.nih.gov/32361528/

